# Effects of a 24-week resistance exercise program on brain amyloid and Alzheimer’s disease blood-based biomarkers: the AGUEDA randomized controlled trial

**DOI:** 10.64898/2026.03.02.26347392

**Authors:** Patricio Solis-Urra, Marcos Olvera-Rojas, Yolanda García-Rivero, Xuemei Zeng, Yijun Chen, Anuradha Sehrawat, Mahnaz Shekari, Lauren E. Oberlin, Kirk I. Erickson, Thomas K. Karikari, Manuel Gómez-Río, Francisco B. Ortega, Irene Esteban-Cornejo

**Author notes:** Corresponding authors: Patricio Solis-Urra, Marcos Olvera-Rojas, Irene Esteban-Cornejo. Department of Physical and Sports Education, Faculty of Sports Science, University of Granada, Granada, Spain. Carretera de Alfacar, s/n CP: 18071 Granada. AdventHealth Research Institute, Neuroscience, Innovation Tower, 265 E Rollins St, Suite 2100 Orlando, FL 32804, USA. Patricio Solis-Urra and Marcos Olvera-Rojas contributed equally to this work and are co-first authors.

## Abstract

We examined whether a 24-week resistance training program influenced brain amyloid-β (Aβ) and Alzheimer’s Disease (AD)-related blood-based biomarkers. Ninety cognitively normal, physically inactive older adults aged 65–80 years were randomly allocated to a 24-week resistance training program (three ∼60-min supervised sessions/week) or a wait-list control group. Primary analyses assessed exercise-induced changes in brain Aβ (Centiloid values) and plasma ptau217/Aβ1-42 IPMS ratio. Secondary analyses examined ptau217/Aβ42 SIMOA ratio, ptau217, ptau181 and Aβ42/40, as well as potential interactions with sex, age, education, apolipoprotein ε4 (*APOE4*) status, amyloid PET-positivity, and comorbidities. The intervention produced no significant differences on brain Aβ or AD-related blood-based biomarkers (p>0.05) compared to the control group. However, the ptau217/Aβ1–42 IPMS ratio showed a small, non-significant increase in the control group (SMD = 0.162; 95% CI: −0.159 to 0.483) while remaining stable in the exercise group (SMD = 0.01; 95% CI: −0.291 to 0.310) with a similar trend for ptau217/Aβ42 SIMOA. Moderator analyses indicated differential responses by amyloid PET-positivity and *APOE4* status on brain Aβ (p for interaction<0.05), with increases observed in *APOE4* carriers and amyloid PET–positive individuals in the control group, whereas those allocated to the exercise intervention reduced their levels. The specificity observed within our subgroups suggests that resistance exercise may serve as a targeted intervention to modulate AD pathophysiology, raising new questions regarding its broader role in the delay of the disease in vulnerable populations.

## Introduction

Amyloid-β (Aβ) plaque accumulation is a key therapeutic target in Alzheimer’s disease (AD), as its deposition drives neurodegeneration and cognitive decline in older adults ^1^. Aβ positron emission tomography (PET) is a gold-standard biomarker to detect brain Aβ plaques, while the current Food and Drug Administration clearance for the Lumipulse ptau217/Aβ1-42 peripheral plasma ratio assay marks an important step toward widespread implementation in clinical practice. Other AD-related blood-based biomarkers (BBMs; i.e., ptau217, ptau181 and Aβ42/Aβ40 ratio), are being investigated intensively as they capture distinct yet complementary aspects of AD pathology, offering a promising avenue for early diagnosis and monitoring as well as significant potential for drug development and trial enrollment ^2–4^.

While pharmaceutical strategies targeting brain Aβ have been developed to delay cognitive decline, with donanemab and lecanemab achieving near-complete clearance of Aβ plaques in a dose-dependent manner and a moderate slowing of cognitive deterioration ^5–8^, non-pharmaceutical interventions have received comparatively less attention and warrant further consideration ^9,10^. As such, physical exercise impacts brain health across the lifespan ^11^, with evidence from animal studies suggesting its neuroprotective effects by mitigating amyloid accumulation, inflammation, microglial activation and tau hyperphosphorylation mechanisms ^12–17^, yet studies in humans are less consistent ^18^. Specifically, there have been three randomized controlled trials (RCTs) investigating the effects of aerobic exercise on brain Aβ accumulation using PET, but none of these studies have used the plasma ptau217/Aβ42 ratio. For instance, a 52-week exercise intervention did not prevent brain Aβ accumulation (quantified by PET global Standardized Uptake Value Ratio [SUVR]) in amnestic mild cognitive impairment patients ^19^ or cognitively normal older adults ^20^, despite slight improvements in memory and executive function scores ^19^. In addition, 16-weeks of aerobic exercise did not affect cortical Aβ PET SUVR in AD patients ^21^. Therefore, current human evidence does not support the notion that aerobic exercise has a detectable impact on brain Aβ accumulation or AD BBMs. Indeed, aerobic exercise represents only one of multiple exercise modalities, each of which may differentially influence brain pathophysiology ^22^. For instance, resistance exercise, which has been shown to induce beneficial effects on brain structure and cognitive function ^23–25^, could therefore plausibly influence mechanisms and pathways relevant to brain Aβ accumulation and AD-related BBMs. In addition to this, it is well established that certain factors (e.g., *APOE4* status, amyloid PET-positivity and education) increase the risk of AD or are associated with greater pathological burden and should be taken into account when interpreting clinical outcomes ^26,27^.

Accordingly, we conducted a RCT to determine the effects of a 24-week resistance exercise program on brain Aβ accumulation, expressed on the Centiloid scale (CL), and on the plasma ptau217/Aβ1-42 IPMS ratio (where Aβ1-42 was quantified by IPMS) in cognitively normal older adults. The secondary objective was to assess effects on other AD-related BBMs (ptau217/Aβ42 SIMOA [both analytes measured by SIMOA], ptau217, ptau181 and Aβ42/Aβ40 ratio, both IPMS and SIMOA). The third objective was to evaluate whether sex, age, education, *APOE4*, amyloid PET-positivity, and comorbidities (e.g., heart disease, obesity) moderated exercise-related changes in these biomarkers. Based on prior evidence in aerobic exercise, we anticipated minimal or no detectable effects of a 24-week resistance exercise program on brain Aβ accumulation and plasma ptau217/Aβ1–42 IPMS ratio. Consequently, we explored potential trends and the influence of individual moderators. We expected that exercise-related changes in these biomarkers would be more pronounced in individuals at higher risk for AD, including APOE4 carriers, amyloid PET–positive participants, and older individuals.

## Materials and methods

### Trial design and patients

The AGUEDA trial (Clinicaltrials.gov NCT05186090) was a single-center, two-arm, single-blind RCT including a total of 90 cognitively normal older adults from Granada, Spain. The primary endpoint of the AGUEDA trial was to evaluate the impact of a 24-week resistance exercise program on executive function, results of which have been published by Fernandez-Gamez, Solis-Urra et al. 2026 ^25^. The design and baseline characteristics of the AGUEDA trial have been reported previously ^28^. In particular, eligible participants were physically inactive (not participating in any resistance exercise program in the last 6 months and <600 METs/week of moderate-to-vigorous physical activity), free of depressive symptoms and aged 65-80 years with a cognitively normal profile defined as: (1) the Spanish version of the modified Telephone Interview of Cognitive Status (STICS-m) (≥26 points) ^29^, (2) Mini-Mental State Examination (MMSE) (≥25/30) ^30^ and (3) Montreal Cognitive Assessment (MoCA) (<71 years, ≥24/30, 71-75≥22/30, >75, 21/30) ^31^. Detailed information about the study inclusion and exclusion criteria, recruitment and screening procedures is available elsewhere ^28^. All participants provided written informed consent to participate in the trial. The reporting of the results adheres to the Consolidated Standards of Reporting Trials Extension (CONSORT) guidelines (see Supplementary material, **Table S3**). The trial protocol was approved by the Research Ethics Board of the Andalusian Health Service (CEIM/CEI Provincial de Granada; #2317-N-19) on May 25th, 2020. Participants were randomized in a 1:1 allocation and in a blinded fashion to the resistance exercise or the wait-list control group, stratified by sex (male vs female) and age (<72 vs >= 72 years). Randomization was managed via REDCAP by an external researcher (Dr. Veronica Cabanas-Sanchez) to maintain blinding. Blinded assessors conducted outcome assessments at baseline and after the 24-week intervention period. Primary outcomes were brain Aβ assessed by PET (i.e., Centiloid) and the ptau217/Aβ1-42 IPMS ratio. Secondary outcomes were other AD-related BBMs (ptau217/Aβ42 SIMOA, ptau217, ptau181 and Aβ42/Aβ40 ratio, both IPMS and SIMOA), as well as potential interactions with sex, age, education, *APOE4* status, amyloid PET positivity, and comorbidities.

### Intervention protocol

Comprehensive details of the intervention protocol, including exercise modality, prescribed dose, intensity, session attendance, and adherence strategies, are provided in the main effects paper and in the corresponding CERT (Consensus on Exercise Reporting Template) publication guidelines ^25,32^. Briefly, enrolled participants underwent three ∼60-min resistance exercise sessions per week or a wait-list control over 24 weeks. The resistance exercise program consisted of a combination of upper and lower limb exercises using elastic bands and the participant’s own body weight in groups of 4 – 6 participants and was conducted by professional trainers with a bachelor’s degree in Sport and Exercise Sciences. Sessions were completed in a fitness room at the University Institute of Sports and Health (IMUDS) in the city of Granada, Spain.

### Brain Aβ PET

Following Alzheimer’s Disease Neuroimaging Initiative (ADNI) protocols ^33^, each participant underwent a 20-minute PET scan beginning approximately 90 minutes (± 20 minutes) after the intravenous injection of 300 megabecquerels (MBq) ± 20% of [18F] Florbetaben (Neuraceq; Piramal Pharma) by a trained nurse. PET images were collected using a Siemens Biograph-Vision 600 Edge Positron Emission Tomography/Computed Tomography (PET/CT) digital scanner at the Virgen de las Nieves University Hospital, Granada, Spain. The entire cerebellum served as the reference region to calculate whole-brain SUVRs using standard volumes of interest (VOIs). These SUVRs were then converted to the CL scale using a previously validated pipeline following standard procedures ^34^.

In our study, we selected a CL cut-off of 12 to categorize participants into two different groups: amyloid PET-negative participants (CL <12) and amyloid PET-positive participants (CL ≥12)^35^. These thresholds were selected based on evidence from the Salvadó et al (2019) ^36^ and La Joie et al (2019) ^37^ studies, as a cut-off of 12 CL aligns with postmortem measures of AD neuropathology and reliably detects moderate-to-frequent neuritic plaques following Consortium to Establish a Registry for Alzheimer’s Disease (CERAD) procedures ^38^.

### Blood-based biomarkers

Classical AD biomarkers, including ptau217, ptau181, Aβ42, Aβ40, were measured in fasted plasma samples collected from study participants using the SIMOA assay conducted on an HD-X platform (Quanterix, Billerica, MA, USA) at the Department of Psychiatry, University of Pittsburgh, USA. Plasma ptau181 was assessed with the ptau181 V2 Advantage kit and ptau217 with the ALZpath ptau217 assay kit. Plasma Aβ42 and Aβ40 were measured using the N4PE assay. Furthermore, plasma Aβ1-42 and Aβ1-40 levels were quantified using an immunoprecipitation mass spectrometry (IPMS) technique developed at the University of Pittsburgh ^39^. For consistency, the nomenclature throughout the text will be Aβ42 or Aβ40 IPMS to distinguish it from Aβ42 or Aβ40 SIMOA. Further details on blood sample processing and biomarker analysis are provided elsewhere ^35^.

### Moderators

Sex, age, education, *APOE4*, amyloid PET-positivity, and comorbidities, were included as potential moderators. Sex was classified as male or female based on self-reported data regarding sex assigned at birth. Age was categorized into two groups: younger (<72 years) and older (≥72 years), using the median age at baseline as the cut-off, which also corresponded to the randomized allocation of participants into exercise groups. Education was categorized into two groups: low (≤12 years) and high (>12 years). Participants were classified as *APOE4* carriers if they had at least one ε4 allele, while those without an ε4 allele were categorized as non-carriers. A cut-off of 12 CL was used to categorize participants into two different groups: amyloid PET-negative (CL <12) and amyloid PET-positive (CL ≥12) participants. Presence of medical comorbidities was categorized based on number (< 3 vs ≥ 3), including conditions related to hypertension, diabetes, heart disease, obesity, and cholesterol.

### Statistical analysis

The main analysis of the primary outcome was based on the intention-to-treat (ITT) principle, assuming missing data to be missing at random and without imputation. The ITT analysis included all randomized participants (n = 90). Continuous outcomes were analyzed using constrained baseline longitudinal analysis via a linear mixed model. All statistical analyses were performed in R, version, 4.5.0. Constrained linear mixed models (CLMM) with restricted maximum likelihood estimation were implemented using LMMstar and lme4. Estimated marginal means, within-group differences, and between-group differences (Exercise vs. Control) were obtained with emmeans. The intervention effect was estimated as the coefficient of the interaction term, reported with 95% confidence intervals (CIs). The model included fixed effects for time (two levels: baseline and 24 weeks), treatment (coded 0 for both groups at baseline and coded 1, 2 at 24 weeks for control and exercise, respectively), and the unique patient identifier as a random effect ^40^. Subsequently, a moderation analysis was conducted to determine the extent to which the observed treatment effect was moderated by sex, age, education, APOE4, amyloid PET-positivity, and comorbidities. Per protocol (PP) analysis was performed with only participants who met 80% attendance to exercise sessions.

## Results

Clinical and demographic characteristics are presented in **Table S1**. Specifically, the baseline mean brain Aβ (CL value) for the entire sample was 7.06 ± 25.25. Based on a CL cut-off of 12, the prevalence of amyloid PET-positivity was 21%, while the prevalence of *APOE4* carriership was 15%.

ITT analyses revealed that after the 24-week resistance exercise intervention, no significant reductions were observed in brain Aβ or the ptau217/Aβ42 IPMS ratio compared to the control group in the full sample (**Figure 1A**; Centiloid: SMD −0.003; 95% CI −0.10 to 0.18; ptau217/Aβ42 IPMS ratio: SMD −0.152; 95% CI −0.53 to 0.23). These differences correspond to absolute group differences of 0.077 CL units and 0.076 in the ptau217/Aβ42 IPMS ratio (**Table S2)**. However, the ptau217/Aβ42 IPMS ratio showed a small, non-significant increase in the control group (SMD = 0.162; 95% CI: −0.16 to 0.48) while remaining stable in the exercise group (SMD = 0.01; 95% CI: −0.29 to 0.31), with a similar trend for the ptau217/Aβ42 SIMOA ratio (control: SMD = 0.15; 95 % CI: −0.2 to 0.49; exercise: SMD = −0.03; 95 % CI: −0.35 to 0.29). No significant effects were detected for the other biomarkers including ptau217, ptau181, or the Aβ42/Aβ40 ratio, both IPMS and SIMOA (**Figure 2; Figure S1**). These findings were consistent across sensitivity and per-protocol analyses (**Figure S2**)

**Figure 1.**
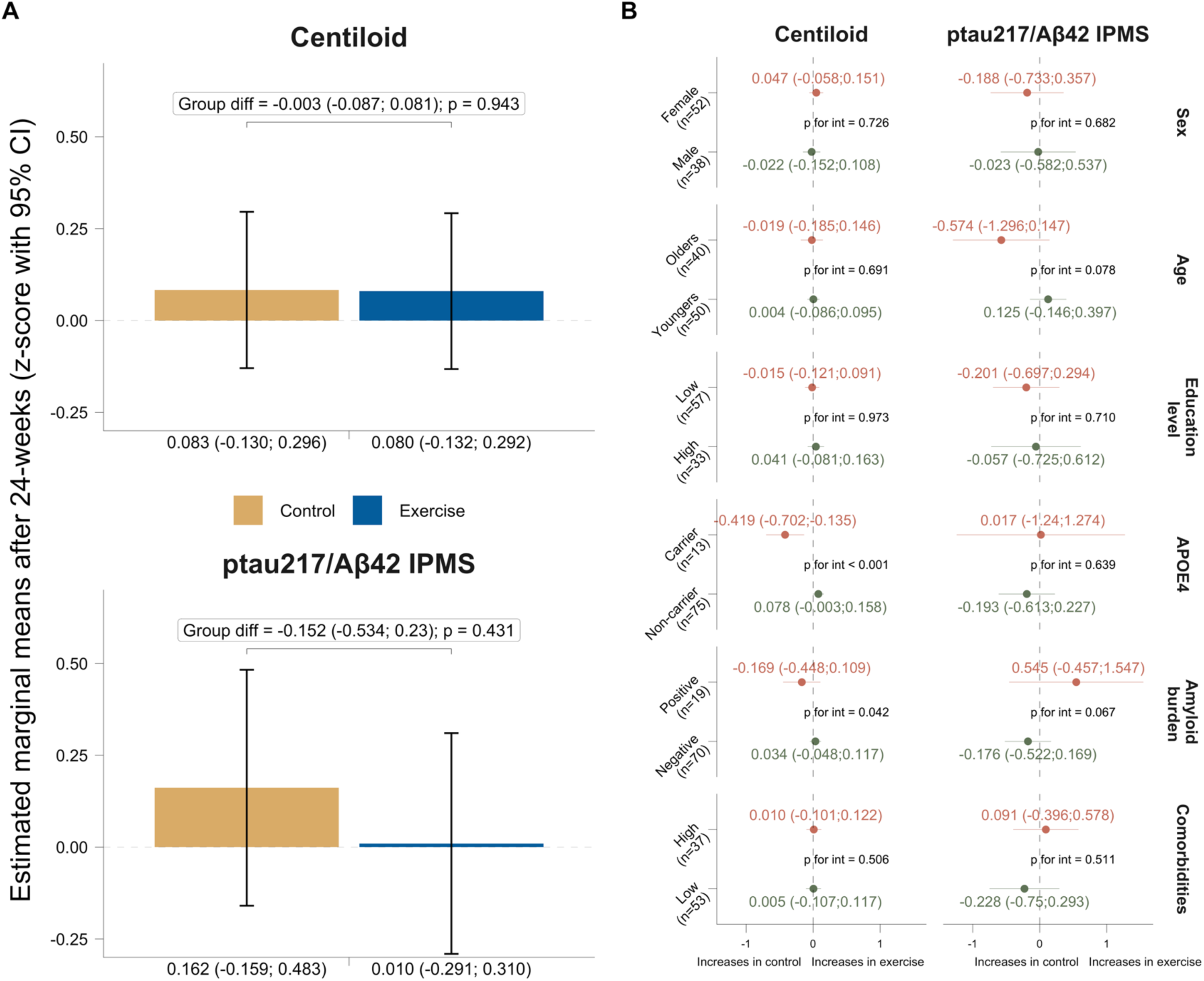
Intention-to-treat effects of the 24-week resistance exercise intervention on Centiloid values and ptau271/Aβ42 ratio in the whole sample (n =90) (A) and according to key moderators (B). Bars indicate marginal means and 95% confidence intervals at 24 weeks for each group (Panel A). Dots represent the mean difference between exercise and control group at 24 weeks (Panel B). As the baseline mean is set to 0, marginal means at 24 weeks can be interpreted as changes in z-score. CI: Confidence interval. IPMS: Immunoprecipitation-mass spectrometry.

**Figure 2.**
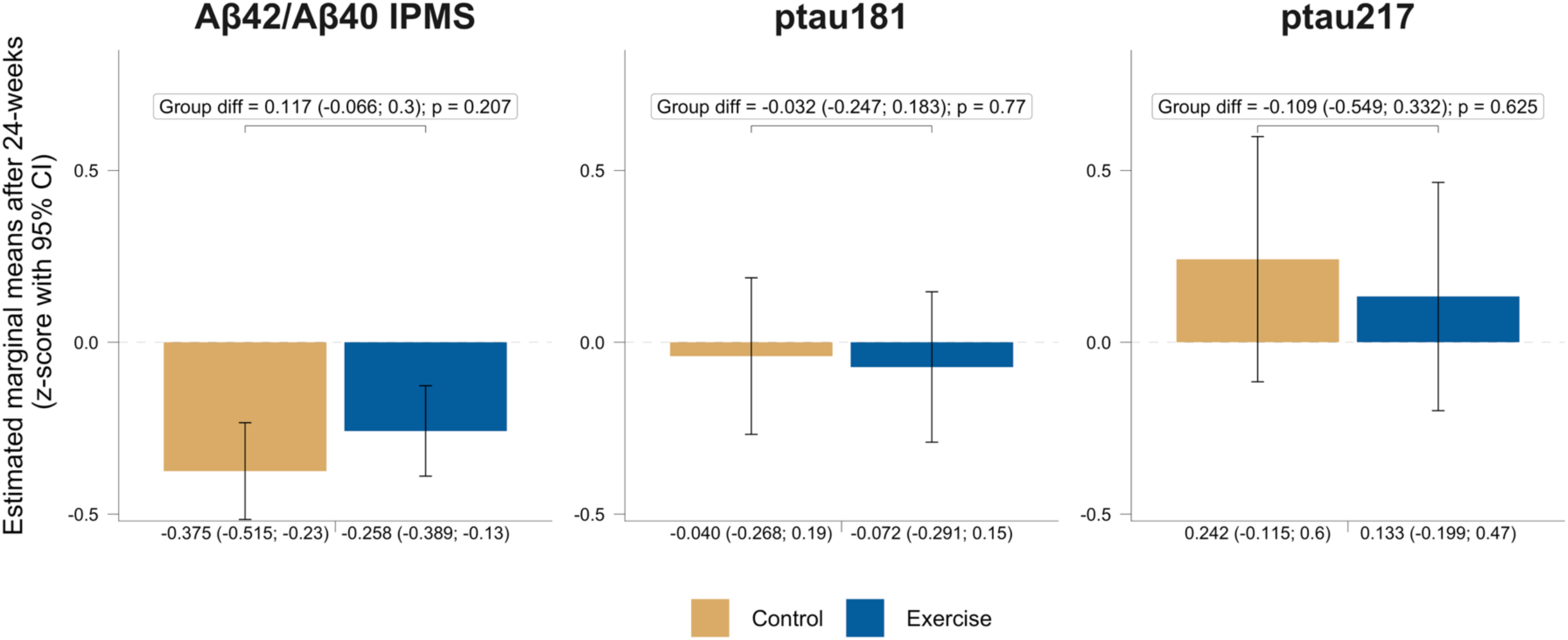
Intention-to-treat effects of the 24-week resistance exercise intervention on other Alzheimer’s disease blood biomarkers in the whole sample (n =90). As the baseline mean is set to 0, marginal means at 24 weeks can be interpreted as changes in z-score. Bars indicate marginal means and 95% confidence intervals at 24 weeks for each group. CI: Confidence interval. IPMS: Immunoprecipitation-mass spectrometry.

Moderator analyses revealed a borderline significant interaction of the intervention with amyloid PET-positivity (p for interaction = 0.042) and with *APOE4* genotype (p for interaction < 0.001) for brain Aβ (**Figure 1B**). While *APOE4* non-carriers and amyloid PET–positive groups remain stable (Figure 3A), increases in brain Aβ values were observed in *APOE4* carriers (Δ CL: 6.875) and amyloid PET–positive participants (Δ CL: 2.847) within the control group (Figure 3B), whereas those assigned to the exercise intervention reduced their levels (*APOE4* carriers, Δ CL: −3.702; amyloid PET–positive: −1.443).

**Figure 3.**
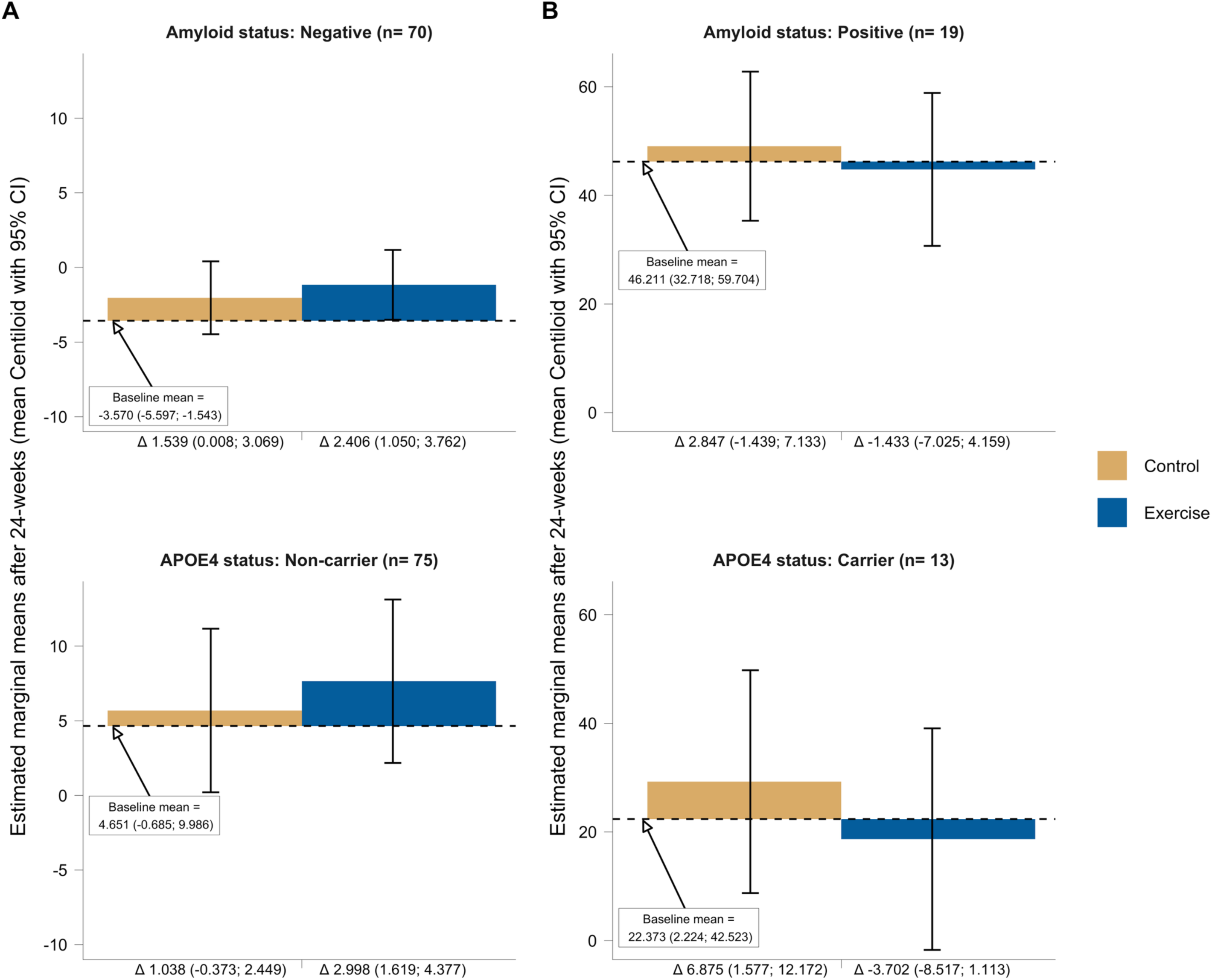
Intention-to-treat effects of the 24-week resistance exercise intervention on additional Alzheimer’s disease blood biomarkers according amyloid-PET and APOE4 status. The dashed horizontal line represents Centiloid values at baseline across both the control and exercise groups. Bars indicate marginal means and 95% confidence intervals at 24 weeks for each group. Values on the x-axis represent the rate of change from the baseline in each group in Centiloid (95% confidence interval). CI: Confidence interval. Δ: Change. Numbers of each analysis: amyloid-PET negative, exercise n= 37, control n= 33; amyloid-PET positive, exercise n= 8, control n= 11; APOE4 carriers, exercise n= 7, control n= 6; APOE4 non-carriers, exercise n= 38, control n= 37.

## Discussion

The main objective of the study was to determine the effects of a 24-week resistance exercise program on brain Aβ and on the plasma ptau217/Aβ42 IPMS ratio in cognitively normal older adults. Overall, we did not identify a significant effect of structured resistance exercise on brain Aβ or the plasma ptau217/Aβ42 IPMS ratio. However, the ptau217/Aβ42 IPMS ratio increased in the control group, whereas it remained stable in the exercise group, with a similar trend for ptau217/Aβ42 SIMOA although these between-group differences did not reach statistical significance. The secondary objective was to assess the effects on other AD-related BBMs (ptau217, ptau181 and Aβ42/Aβ40, both IPMS and SIMOA). We did not observe any significant effect of our intervention on these biomarkers. The third objective was to evaluate whether sex, age, education, *APOE4*, amyloid PET-positivity, and comorbidities moderated exercise-related changes. Results indicated that the effects of the exercise intervention differed according to *APOE4* and amyloid PET-positivity regarding brain Aβ values, with increases observed in *APOE4* carriers and amyloid PET–positive individuals in the control group, whereas those allocated to the exercise intervention reduced their levels.

Our study is one of the first to investigate whether resistance exercise influences brain Aβ accumulation and the ptau217/Aβ42 IPMS ratio in cognitively normal older adults. The null findings regarding brain Aβ accumulation measured by PET are consistent with previous literature in aerobic exercise interventions ^19–21^. These null findings suggest that the duration of the interventions in humans may have been insufficient to replicate the timeframes used in animal studies ^41^. What is more, the greater diversity in disease progression, the lack of controlled conditions in humans compared to animals, and the superior granularity of data obtained from invasive methods such as brain dissection in animal studies, which surpass the resolution and detail achievable through non-invasive imaging in humans, could also explain the lack of effect detection ^42–44^. Considering that brain Aβ plaques may take several decades to accumulate ^45^, it is plausible that 24 to 52-weeks of exercise interventions may be insufficient to produce detectable changes in the brain Aβ burden in the overall population. Nonetheless, it is possible that individuals at higher risk, such as *APOE4* carriers or amyloid-positive participants, could exhibit more readily detectable changes, as these individuals often have faster baseline rates of amyloid accumulation and higher levels of existing pathology, which may increase the likelihood of observing measurable intervention effects.

Indeed, we found two significant moderations for brain Aβ, involving amyloid PET-positivity and *APOE4* carriership. Specifically, in high-risk subgroups, including *APOE4* carriers and amyloid PET–positive individuals, increases in brain Aβ were observed among those in the control group, whereas no significant reductions were detected in the exercise group. Because individuals with established amyloid pathology (amyloid-positive individuals) and increased genetic risk (*APOE4* carriers) typically exhibit faster baseline rates of amyloid accumulation, longitudinal increases, higher than those typically observed in the overall population (∼ 3CL/year), are expected in the absence of an active intervention ^27,46,47^. Our amyloid-positive individuals show a rate of increase of ∼3 CL after 24-weeks, comparing to the ∼1.5 CL rate of increase of negative individuals. This higher rate of change also makes it easier to detect potential effects of interventions, such as the stabilization observed in the exercise group (decrease of ∼ 1.4 CL in 24 weeks). Although these interactions reached statistical significance, they should be interpreted with caution given the relatively small sample size available for these subgroup analyses. Therefore, these findings should be considered exploratory and warrant replication in larger, independent cohorts to confirm their robustness.

On the other hand, peripheral AD-related blood-based biomarkers may represent more sensitive proxies to capture early or dynamic biological responses to lifestyle-based interventions such as exercise ^48^. To note, our study is among the first to examine the effects of resistance exercise on AD-related BBMs such as ptau217/Aβ42 IPMS, ptau217/Aβ42 SIMOA and ptau217. Although no significant differences emerged between the control and exercise groups, the resistance exercise group appeared to maintain their biomarker levels, particularly for the ptau217/Aβ42 IPMS ratio, whereas the control group showed a tendency toward an increase, with a similar trend for ptau217/Aβ42 SIMOA. This finding is particularly relevant given the growing importance of the ptau217/Aβ42 ratio ^49^, which has emerged as one of the most accurate blood-based predictors of brain amyloid plaques ^35,50^. Based on the consistent stabilization pattern observed for ptau217/Aβ42 across platforms (IPMS and SIMOA), the slow temporal dynamics of brain amyloid in humans ^51^, and evidence from long-duration exercise interventions in animal models ^52^, it is therefore reasonable to hypothesize that a longer intervention period might have reinforced or extended the observed pattern in the ptau217/Aβ42 ratio. Considering the dose-dependent nature of resistance exercise effects ^53^, and that the exercise was performed at a mean rating of perceived exertion of 5.4 on a 0–10 scale ^25^, corresponding to a moderate-to-low relative intensity, it remains possible that delivering higher-intensity resistance exercise over a 24-week period, or longer, could yield more pronounced effects on this blood-based biomarker. Thus, the AGUEDA program was able to stabilize biomarker trajectories yet may have provided insufficient peripheral stimulus to drive more robust systemic signaling. Similarly, no significant group differences were observed for other AD-related BBMs (i.e., ptau217, ptau181 and Aβ42/Aβ40, both IPMS and SIMOA). Future trials with extended intervention durations and higher-intensity or progressively externally-overloaded exercise stimuli, combined with sufficiently powered designs, will be essential to determine whether resistance training can meaningfully modulate or stabilize the longitudinal trajectories of these blood biomarkers.

A limitation of our study is that statistical power was calculated based on the trial’s primary outcome, and the present analysis represents a secondary investigation. This may have reduced our ability to detect meaningful changes in the outcomes examined, potentially constraining the observable impact of the intervention on brain Aβ and the ptau217/Aβ42 ratios. Future trials should consider incorporating these biomarkers as primary endpoints and enrolling populations at higher risk of AD, with a priori statistical power and sample size calculations specifically tailored to these outcomes, to more robustly evaluate the exercise-related effects and the interactions suggested by our findings. Nevertheless, our study is among the first to examine the effects of a structured resistance training program on both central and peripheral markers, including the ptau217/Aβ42 ratios measured by IPMS and SIMOA, as well as ptau217 itself. Future research should investigate whether higher-intensity or longer-duration interventions can elicit more pronounced effects, and whether combining exercise with additional non-pharmacological or pharmacological strategies produces additive or synergistic benefits.

In conclusion, the results of this trial indicate that 24 weeks of resistance exercise did not produce significant changes in brain Aβ or AD-related blood-based biomarkers in the overall sample of cognitively normal older adults. However, subgroup analyses revealed that brain Aβ increased in vulnerable participants (i.e., APOE4 carriers and amyloid PET–positive) in the control group, whereas those assigned to the exercise intervention reduced their levels. Moreover, while group differences did not reach significance, the apparent stabilization of ptau217/Aβ42 ratios in the exercise group, while increasing in the control group, hints that longer or more demanding interventions might strengthen these effects. Overall, these results highlight gaps in our understanding of resistance exercise and raise new questions about its potential role in AD prevention and delay.

## Data Availability

The datasets generated and/or analyzed during the current study are not publicly available due to ethical and institutional restrictions but are available from the corresponding authors on reasonable request.

## Supplementary material

**Figure S1.**
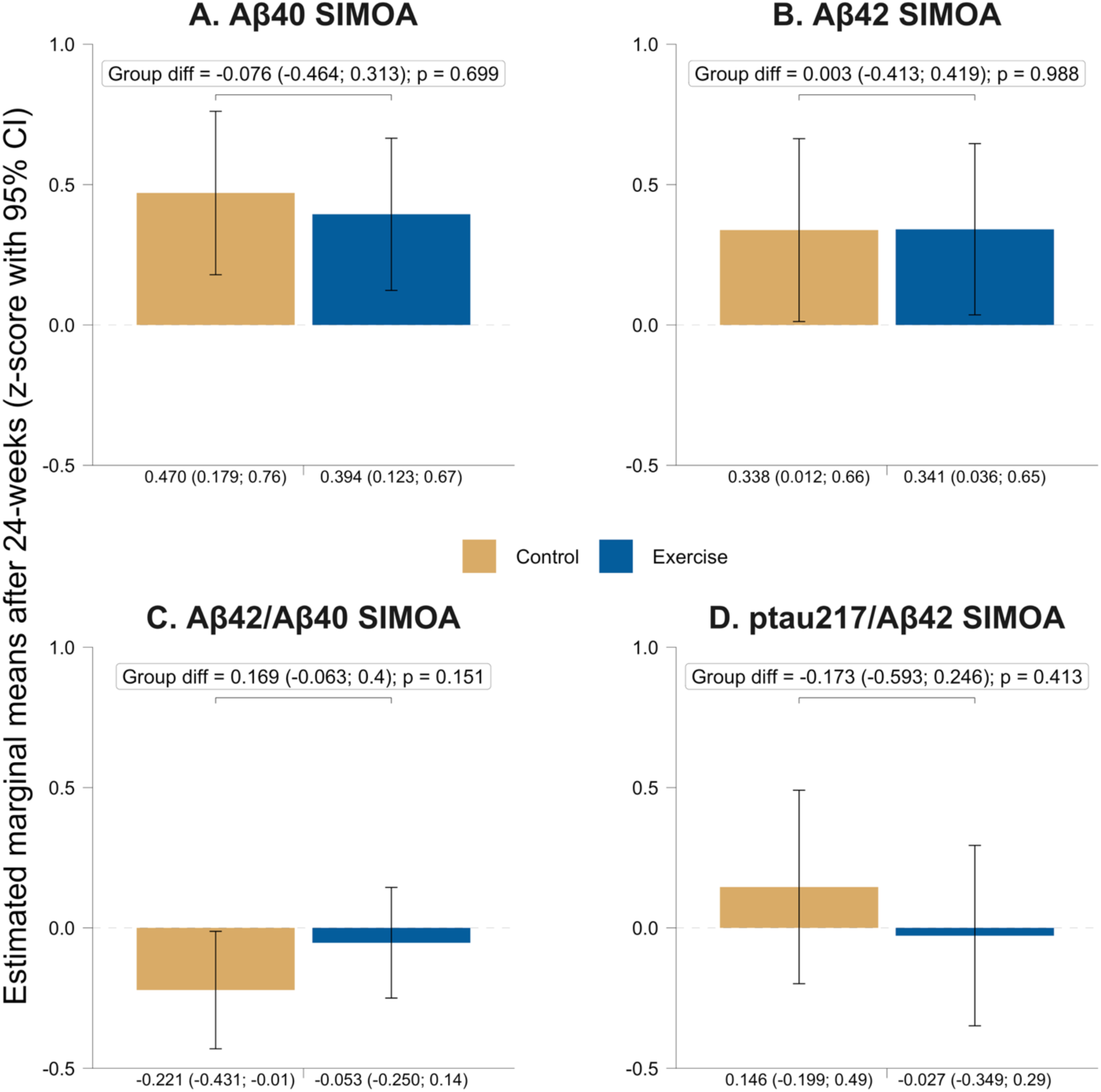
Intention-to-treat effects of the 24-week resistance exercise intervention on AD blood biomarkers using SIMOA in whole sample (n =90). Bars indicate marginal means and 95% Confidence Interval at 24 weeks in each subgroup. CI: Confidence interval. As the baseline mean is set to 0, marginal means at 24 weeks can be interpreted as changes in z-score.

**Figure S2.**
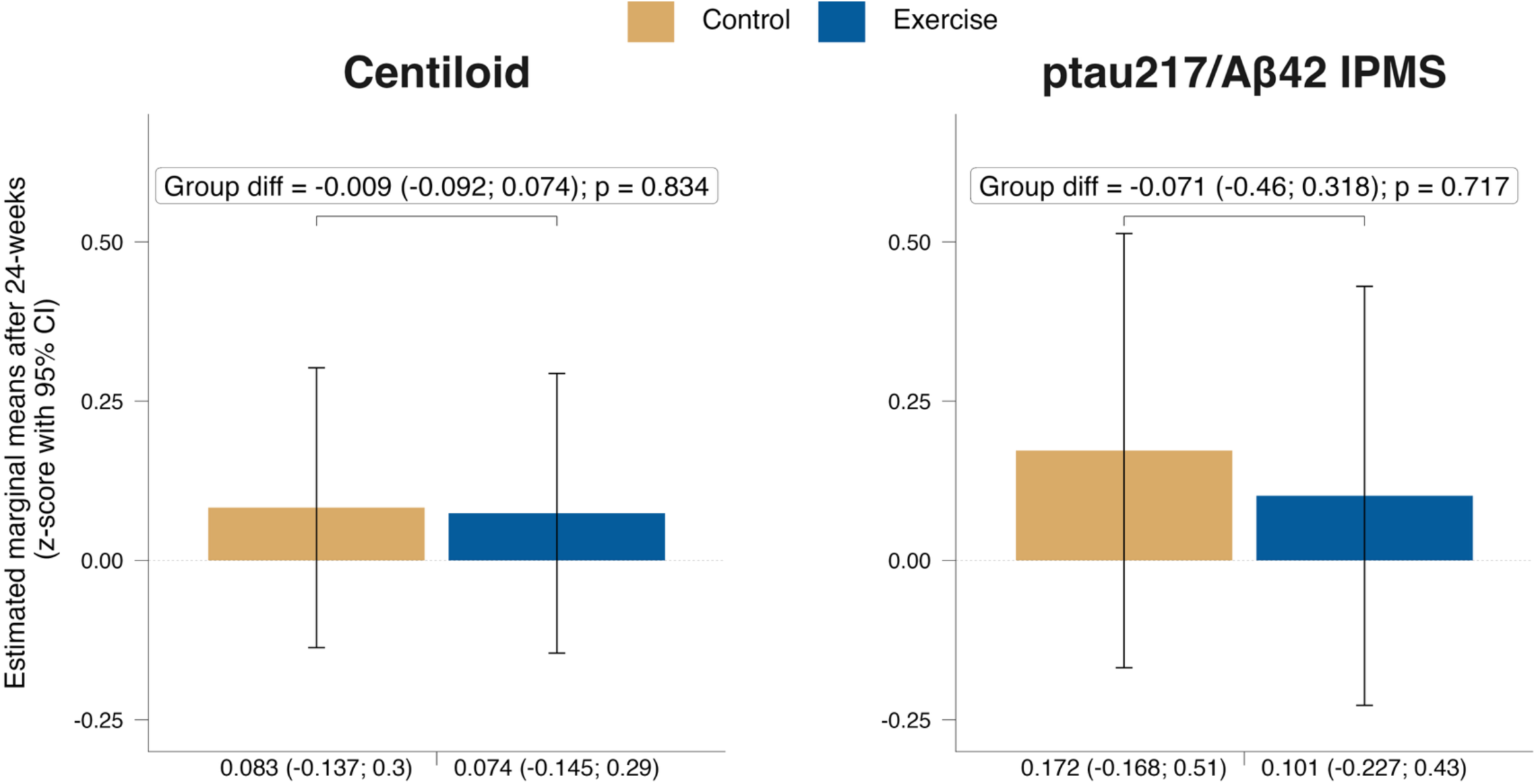
Per protocol effects of the 24-week resistance exercise intervention on Centiloid and ptau217/Aβ42 IPMS ratio (n=84). Bars indicate marginal means and 95% Confidence Interval at 24 weeks in each subgroup. As the baseline mean is set to 0, marginal means at 24 weeks can be interpreted as changes in z-score. CI: Confidence interval.

**Table S1.** Baseline descriptive characteristics of the AGUEDA sample.

| Characteristic | N | Overall<br>N = 90 | Control<br>N = 44 | Exercise<br>N = 46 |
| --- | --- | --- | --- | --- |
| Sex, Female, n (%) | 90 | 52 (58%) | 25 (57%) | 27 (59%) |
| Age at baseline, yr, | 90 | 71.75 ± 3.96 | 71.58 ± 3.73 | 71.90 ± 4.21 |
| Education length, yr, | 90 | 11.54 ± 4.90 | 11.93 ± 4.53 | 11.17 ± 5.26 |
| Education level (> 12 years), n (%) | 90 | 33 (37%) | 19 (43%) | 14 (30%) |
| Body-mass index, kg/m<sup>2</sup>, | 90 | 28.50 ± 4.23 | 28.67 ± 4.05 | 28.35 ± 4.43 |
| APOE4 carrier, n (%) | 88 | 13 (15%) | 6 (14%) | 7 (16%) |
| Amyloid PET-positive, n (%) | 89 | 19 (21%) | 11 (25%) | 8 (18%) |
| Subjective cognitive decline, score, | 90 | 2.92 ± 1.97 | 3.11 ± 1.94 | 2.74 ± 1.99 |
| Telephone Interview of Cognitive Status, score, | 90 | 33.91 ± 2.72 | 34.05 ± 2.81 | 33.78 ± 2.65 |
| Mini-Mental State Examination, score, | 90 | 28.92 ± 1.06 | 28.84 ± 1.16 | 29.00 ± 0.97 |
| Montreal Cognitive Assessment, score, | 90 | 25.58 ± 2.14 | 25.89 ± 2.05 | 25.28 ± 2.21 |
| Centiloid, | 89 | 7.06 ± 25.25 | 10.33 ± 26.78 | 3.86 ± 23.52 |
| ptau217/Aβ42 IPMS, | 89 | 0.96 ± 0.50 | 0.95 ± 0.45 | 0.96 ± 0.56 |
| ptau217, pg/ml, | 89 | 0.30 ± 0.15 | 0.30 ± 0.13 | 0.31 ± 0.16 |
| ptau181, pg/ml, | 90 | 2.60 ± 1.33 | 2.50 ± 1.34 | 2.69 ± 1.32 |
| Aβ42/Aβ40, IPMS, | 90 | 0.12 ± 0.02 | 0.12 ± 0.03 | 0.12 ± 0.02 |
| Aβ42/Aβ40, SIMOA, | 88 | 0.06 ± 0.01 | 0.06 ± 0.01 | 0.06 ± 0.01 |
| ptau217/Aβ42 SIMOA, | 87 | 0.06 ± 0.03 | 0.06 ± 0.03 | 0.06 ± 0.04 |

Note: Unless otherwise specified, data are presented as mean ± standard deviation. Abbreviations: m, meters; n, number; SD, standard deviation; sec, seconds; yr, years; AD, Alzheimer disease; IPMS, Immunoprecipitation Mass Spectrometry; SIMOA, Single-molecule array.

**Table S2.** Intention-to-treat effects of the 24-weeks resistance exercise intervention on AD biomarkers.

|  | Estimated marginal means from each timepoint 24-weeks |  |  |  |  |
| --- | --- | --- | --- | --- | --- |
|  | Baseline, Whole sample | Exercise | Control | Group diff | p value |
| Centiloid | 7.251 (1.975;12.527) | 9.078 (3.723;14.434) | 9.155 (3.781;14.529) | -0.077 (-2.196;2.043) | 0.943 |
| ptau217/Aβ42 IPMS | 0.959 (0.854;1.065) | 0.964 (0.814;1.115) | 1.041 (0.879;1.202) | -0.076 (-0.268;0.116) | 0.431 |
| ptau217 | 0.305 (0.274;0.336) | 0.325 (0.276;0.373) | 0.341 (0.288;0.393) | -0.016 (-0.081;0.049) | 0.625 |
| ptau181 | 2.595 (2.317;2.873) | 2.5 (2.209;2.79) | 2.542 (2.239;2.844) | -0.042 (-0.328;0.244) | 0.77 |
| Aβ42/Aβ40 IPMS | 0.121 (0.116;0.126) | 0.115 (0.112;0.118) | 0.112 (0.109;0.116) | 0.003 (-0.002;0.007) | 0.207 |
| ptau217/Aβ42 SIMOA | 0.06 (0.053;0.068) | 0.059 (0.048;0.069) | 0.065 (0.053;0.076) | -0.006 (-0.019;0.008) | 0.413 |
| Aβ42 SIMOA | 5.349 (5.088;5.609) | 5.775 (5.4;6.15) | 5.771 (5.371;6.172) | 0.004 (-0.507;0.515) | 0.988 |
| Aβ40 SIMOA | 91.513 (87.208;95.817) | 99.638 (94.095;105.18) | 101.188 (95.235;107.14) | -1.55 (-9.494;6.394) | 0.699 |
| Aβ42/Aβ40 SIMOA | 0.059 (0.056;0.062) | 0.058 (0.056;0.061) | 0.056 (0.054;0.059) | 0.002 (-0.001;0.005) | 0.151 |

Abbreviations: SD, standard deviation; IPMS, Immunoprecipitation Mass Spectrometry; SIMOA, Single-molecule array. p-tau, phosphorylated tau. Values indicated estimated marginal means (and 95% Confidence Intervals) after 24 weeks of exercise intervention.

**Table S3.** CONSORT (Consolidated Standards of Reporting Trials) checklist.

| Section / Topic | Item No. | CONSORT 2025 checklist item | Reported on Page No |
| --- | --- | --- | --- |
| **Title and abstract** |  |  |  |
| Title and structured abstract | 1a | Identification as a randomized trial | 0 |
|  | 1b | Structured summary of the trial design, methods, results, and conclusions | 0 |
| **Open science** |  |  |  |
| Trial registration | 2 | Name of trial registry, identifying number (with URL) and date of registration | 7 |
| Protocol and statistical analysis plan | 3 | Where the trial protocol and statistical analysis plan can be accessed | 7 |
| Data sharing | 4 | Where and how the individual de-identified participant data (including data dictionary), statistical code and any other materials can be accessed | 3 |
| Funding and conflicts of interest | 5a | Sources of funding and other support (e.g., supply of drugs), and role of funders in the design, conduct, analysis and reporting of the trial | 2 |
|  | 5b | Financial and other conflicts of interest of the manuscript authors | 3 |
| **Introduction** |  |  |  |
| Background and rationale | 6 | Scientific background and rationale | 5 |
| Objectives | 7 | Specific objectives related to benefits and harms | 6 |
| **Methods** |  |  |  |
| Patient and public involvement | 8 | Details of patient or public involvement in the design, conduct and reporting of the trial | - |
| Trial design | 9 | Description of trial design including type of trial (e.g., parallel group, crossover), allocation ratio, and framework (e.g., superiority, equivalence, non-inferiority, exploratory) | 7 |
| Changes to trial protocol | 10 | Important changes to the trial after it commenced including any outcomes or analyses that were not prespecified, with reason | 7 |
| Trial setting | 11 | Settings (e.g., community, hospital) and locations (e.g., countries, sites) where the trial was conducted | 7 |
| Eligibility criteria | 12a | Eligibility criteria for participants | 7 |
|  | 12b | If applicable, eligibility criteria for sites and for individuals delivering the interventions (e.g., surgeons, physiotherapists) | - |
| Intervention and comparator | 13 | Intervention and comparator with sufficient details to allow replication. If relevant, where additional materials describing the intervention and comparator (e.g., intervention manual) can be accessed | 8 |
| Outcomes | 14 | Pre-specified primary and secondary outcomes, including the specific measurement variable (e.g., systolic blood pressure), analysis metric (e.g., change from baseline, final value, time to event), method of aggregation (e.g., median, proportion), and time point for each outcome | 7 |
| Harms | 15 | How harms were defined and assessed (e.g., systematically, non-systematically) | 8 |
| Sample size | 16a | How sample size was determined, including all assumptions supporting the sample size calculation | Secondary outcome within the trial |
|  | 16b | Explanation of any interim analyses and stopping guidelines | - |
| **Randomization:** |  |  |  |
|  | 17a | Who generated the random allocation sequence and the method used | 7 |
| Sequence generation | 17b | Type of randomization and details of any restriction (e.g., stratification, blocking and block size) | 7 |
| Allocation concealment mechanism | 18 | Mechanism used to implement the random allocation sequence (e.g., central computer/telephone; sequentially numbered, opaque, sealed containers), describing any steps to conceal the sequence until interventions were assigned | 7 |
| Implementation | 19 | Whether the personnel who enrolled and those who assigned participants to the interventions had access to the random allocation sequence | 7 |
| Blinding | 20a | Who was blinded after assignment to interventions (e.g., participants, care providers, outcome assessors, data analysts) | 7 |
|  | 20b | If blinded, how blinding was achieved and description of the similarity of interventions | 7 |
| Statistical methods | 21a | Statistical methods used to compare groups for primary and secondary outcomes, including harms | 10-11 |
|  | 21b | Definition of who is included in each analysis (e.g., all randomized participants), and in which group | 10-11 |
|  | 21c | How missing data were handled in the analysis | 10-11 |
|  | 21d | Methods for any additional analyses (e.g., subgroup and sensitivity analyses), distinguishing prespecified from post-hoc | 10-11 |
| **Results** |  |  |  |
| Participant flow, including flow diagram | 22a | For each group, the numbers of participants who were randomly assigned, received intended intervention, and were analysed for the primary outcome | Fernandez-Gamez et al, 2026 |
|  | 22b | For each group, losses and exclusions after randomization, together with reasons | Fernandez-Gamez et al, 2026 |
| Recruitment | 23a | Dates defining the periods of recruitment and follow-up for outcomes of benefits and harms | 7 |
|  | 23b | If relevant, why the trial ended or was stopped | - |
| Intervention and comparator delivery | 24a | Intervention and comparator as they were actually administered (e.g., where appropriate, who delivered the intervention/comparator, whether participants adhered, whether they were delivered as intended [fidelity]) | 7 |
|  | 24b | Concomitant care received during the trial for each group | - |
| Baseline data | 25 | A table showing baseline demographic and clinical characteristics for each group | Table 1 |
| Numbers analysed, outcomes and estimation | 26 | For each primary and secondary outcome, by group:<br>the number of participants included in the analysis<br>the number of participants with available data at the outcome time point<br>result for each group, and the estimated effect size and its precision (such as 95% confidence interval)<br>for binary outcomes, presentation of both absolute and relative effect size | Fernandez-Gamez et al, 2026 |
| Harms | 27 | All harms or unintended events in each group | 7 |
| Ancillary analyses | 28 | Any other analyses performed, including subgroup and sensitivity analyses, distinguishing pre-specified from post-hoc | 10-11 |
| **Discussion** |  |  |  |
| Interpretation | 29 | Interpretation consistent with results, balancing benefits and harms, and considering other relevant evidence | 12-13-14 |
| Limitations | 30 | Trial limitations, addressing sources of potential bias, imprecision, generalisability, and, if relevant, multiplicity of analyses | 12-13-14 |

